# Co-relation of Quadriceps Muscle index (QMI) with Hand Grip Strength (HGS) and Model for End Stage Liver Disease (MELD) score and cut offs for predicting sarcopenia in patients with cirrhosis: A Cross-Sectional study

**DOI:** 10.1101/2025.05.22.25328126

**Authors:** Sarthak Gaur, Minakshi Dhar, Anand Sharma, Sonal Saran, Mukesh Chand Bairwa, Monika Pathania

## Abstract

**Background:** Sarcopenia is a frequent complication of cirrhosis and is closely linked with adverse medical consequences. This study evaluated relationship between the Quadriceps Muscle Index (QMI), measured via ultrasound, and the Model for End-Stage Liver Disease (MELD) score, as well as handgrip strength (HGS) and determine the cut off of QMI that will predict sarcopenia.

**Methodology:** The cross-sectional observational study was carried out at a tertiary care teaching hospital from northern India. Patients having cirrhosis were screened based on predefined eligibility criteria. The assessment followed a multi-step approach, including cirrhosis severity evaluation using the MELD score, muscle mass measurement via the ultrasound-derived Quadriceps Muscle Index (QMI), and muscle strength assessment using hand grip dynamometry and sarcopenia diagnosis using European Working Group on Sarcopenia in Older people 2 (EWGSOP 2) criteria.

**Results:** The study involved 102 patients with cirrhosis. The correlation analysis found moderate negative association among the MELD score and QMI (Spearman rho = - 0.55, p < 0.001). Similarly, substantial positive connection was reported between HGS and QMI (Pearson r = 0.63, p < 0.001). At a cutoff of QMI (cm/m2) ≤1.65 cm/m^2^ for males and ≤1.59 cm/m^2^ for females found to predict sarcopenia in cirrhotic patients.

**Conclusions:** Present study highlights the Quadriceps Muscle Index (QMI) as a reliable and clinically relevant tool for assessing sarcopenia in cirrhotic patients. The significant correlation between QMI and the MELD score underscores its potential for identifying patients at risk of complications of cirrhosis.

## INTRODUCTION

Sarcopenia has been characterized by the slow loss of muscular mass, strength, and function and is increasingly recognized as a crucial determinant of severe health effects, which includes disablement, fractures, plus prolonged stay in the hospital. [1-3] It is categorized into primary sarcopenia, which occurs with aging, and secondary sarcopenia, which results from chronic conditions such as cirrhosis, heart failure, and CKD. [4,5] Sarcopenia in patients having cirrhosis leads to poor outcomes, such as a higher risk of infections, variceal bleeding, hepatic encephalopathy, and unfavourable post-transplant prognosis.[6]

Need for early diagnosis of sarcopenia in patients with cirrhosis is undebatable. European Working Group on Sarcopenia in Older people 2 (EWGSOP 2) defines sarcopenia as-Probable sarcopenia (low muscle strength), Confirmed sarcopenia (low muscle strength and low muscle quality/quantity) and Severe sarcopenia (low muscle strength, low muscle quantity/quality and impaired physical performance). [7] Muscle strength is measured by assessing hand grip strength and muscle performance is assessed by assessing gait speed. The most accurate diagnostic modalities for assessing muscle mass are Magnetic resonance imaging (MRI) and Computed tomography (CT). However, they are costly, time-consuming, and require exposure to radiation. [8] Dual-energy X-ray absorptiometry (DEXA) is extremely reliable way to determine muscle mass.; however, its availability is limited. [9] Bioelectrical impedance analysis (BIA), while accessible, is affected by hydration status and ascites, limiting its accuracy in cirrhotic patients. [9]

Ultrasound has emerged as a promising, cost-effective, and bedside modality for assessing muscle mass, making it a valuable tool for detecting sarcopenia. [1,8] Muscle mass in our study was measured using ultrasound-derived Quadriceps Muscle Index (QMI).

The purpose of this study was to determine the co-relationship between QMI, as determined by ultrasound, with the MELD score and QMI among cirrhosis patients. Also an attempt was made to determine the cut off of QMI that will predict sarcopenia per EWGSOP 2 criteria. Early identification of muscle loss using ultrasound could allow for timely nutritional and rehabilitative interventions, ultimately improving risk stratification, treatment planning, and transplant outcomes in cirrhotic patients.

## METHODOLOGY

A prospective observational cross-sectional study was undertaken in the outpatient departments for General Medicine, Geriatric Medicine, and Gastroenterology at a tertiary centre of North India. Data collection was carried out over an 18-month period. Before the study began, the institutional ethics committee approved it, and all participants gave their informed consent in compliance with ethical standards. Patients aged 18 and older with a definitive diagnosis of cirrhosis, established through radiological, endoscopic, histological, or clinical criteria were used for the study. Patients having malignancies, including hepatocellular carcinoma, Organ failure affecting nutritional well-being (for example, CKD necessitating haemodialysis, heart failure, or respiratory impairment), Acute-on-chronic liver failure (ACLF) or acute decompensation, HIV or tuberculosis infection, diabetes, neurological or musculoskeletal conditions preventing muscle strength or function assessment, chronic pancreatitis were omitted.

### Sample Size Calculation

To ensure statistical robustness and reproducibility, the sample size was estimated using data from a recently published study, which found a correlation coefficient of 0.53 between Quadriceps Muscle Index (QMI) and MELD scores and 0.82 between QMI and HGS in patients with cirrhosis. Assuming a statistical power of 80% and a 5% alpha error, the required sample size for achieving statistical significance was determined to be 102 participants. [8]

### Data Collection

Detailed data on demographic and clinical characteristics were collected from all consecutive participants, including medical history, cirrhosis aetiology, and MELD score calculations based on serum bilirubin, creatinine, and INR values.

**Muscle strength** was assessed using the JAMAR dynamometer. Participants were instructed to apply maximum force while gripping the device, with each hand tested 3 times. The maximum recorded value was used for muscle strength assessment. Low muscle strength taken < 27 kg for males and <16 Kg for females as per EWGSOP 2 criteria.[7]

**Muscle mass** was assessed using the ultrasound-based Quadriceps Muscle Index (QMI). Participants were positioned supine lower limbs relaxed and flexed, while upper limbs supinated. Muscle layer thickness was measured using **B-mode HDI-5000 ultrasound** machine with a broad-band linear array transducer and at a frequency of 5–7.5 MHz. The anterior mid-thigh point was determined as the intersection of the greater trochanter and lateral knee joint line, and right and left thigh thickness of the quadriceps was also taken. [8] QMI = (Quadriceps thickness (Right) + Quadriceps thickness (Left)) / Height^2^ (m^2^)

**Physical performance** was assessed through gait speed. Gait speed (GS) of ≤ 0.8 m/s was used to define reduced physical performance. [7]

### Statistical Analysis

Pearson’s correlation was used for normally distributed variables, whereas Spearman’s correlation was used for non-normally distributed variables. Statistical analysis was performed using SPSS version 23, with p-values <0.05 indicating significance.

## RESULTS

A total of 102 cirrhotic patients were selected from 208 screened according to predetermined criteria in the study. **(Study flow Figure 1 supplemental file)** Table 1 (**supplemental file)** shows the demographic and clinical characteristics of the study population. 45.56 ± 11.08 years was the mean age with most (32.4%) aged 41–50. Males comprised 74.5% of the sample. Alcohol use (46.1%) was most common cause for cirrhosis which was followed by hepatitis C virus (30.4%). Most patients (77.5%) had decompensated cirrhosis.

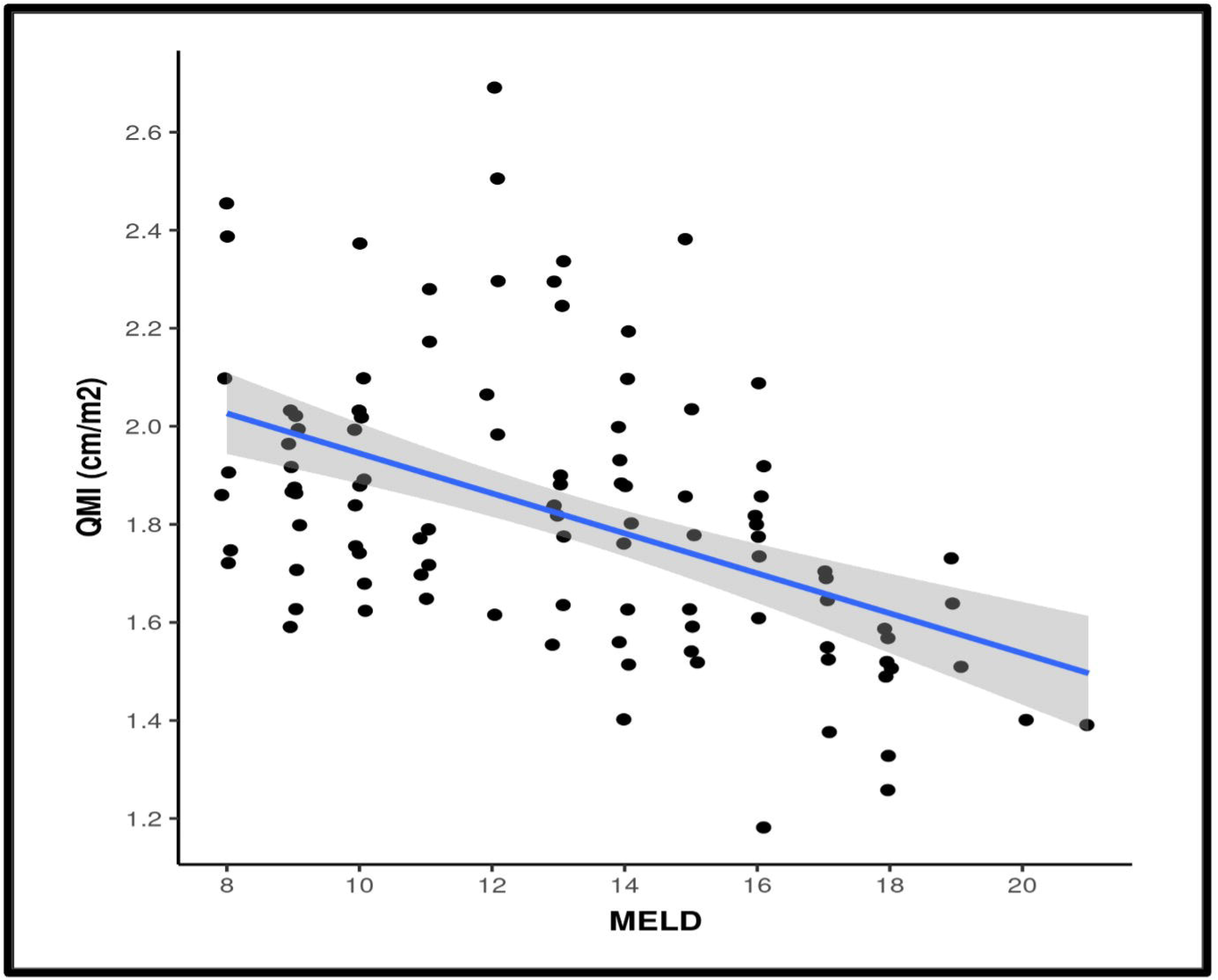

**Table 1:** Correlation between ‘Quadriceps muscle index’ (QMI) and MELD score and ‘Hand grip strength’ and QMI.

| Correlation | Correlation Coefficient | P Value |
| --- | --- | --- |
| MELD vs QMI (cm/m<sup>2</sup>) | -0.55<br>(95%CI: -0.68 to -0.39)<br>(Spearman) | <0.001 |
| HGS vs QMI (cm/m<sup>2</sup>) | 0.63<br>(95%CI: 0.49 to 0.73)<br>(Pearson's) | <0.001 |

A moderate negative correlation (ρ = −0.55) between MELD and QMI (cm/m^2^) was observed, which was statistically significant (p < 0.001) (Table 2). Spearman’s test was used as MELD was not normally distributed. Figure 2 showing scatterplot illustrates the correlation between MELD score and QMI (cm/m^2^), with a blue trendline indicating the relationship and a shaded grey area representing its 95% confidence interval. For every 1 unit increase in MELD, the QMI (cm/m^2^) decreases by 0.04 units.

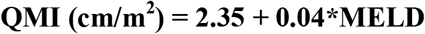

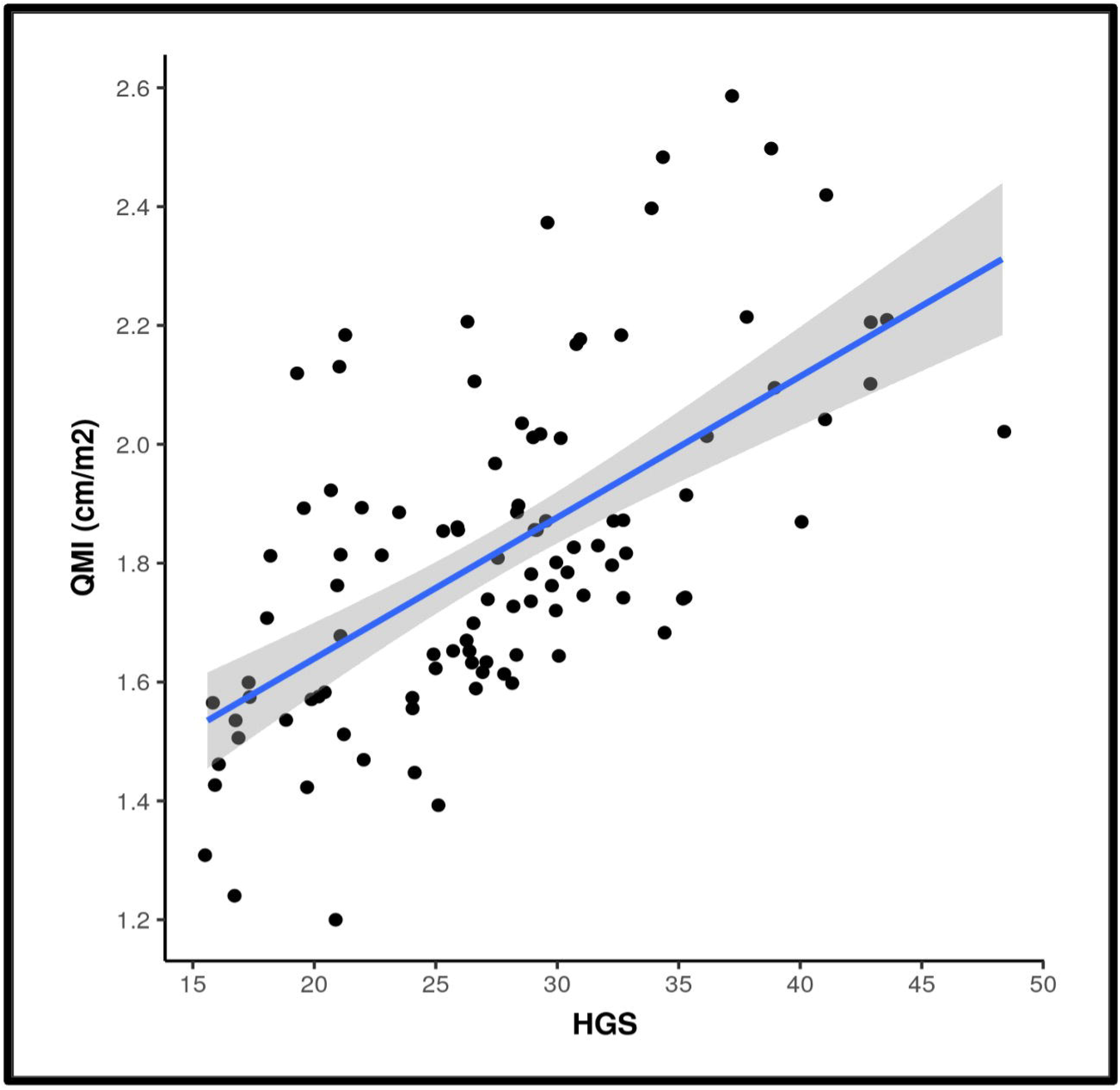

A positive correlation (r = 0.63) was found among HGS and QMI (cm/m^2^) using Pearson’s test, which was statistically significant (p < 0.001), with both variables following a normal distribution. (Table 2) For every 1 unit increase in HGS, the QMI (cm/m2) increases by 0.02 units.

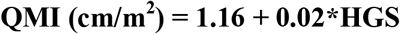

Figure 3 showing scatterplot illustrates the correlation between HGS and QMI (cm/m^2^), with a blue trendline indicating the relationship and a shaded grey area representing its 95% confidence interval.

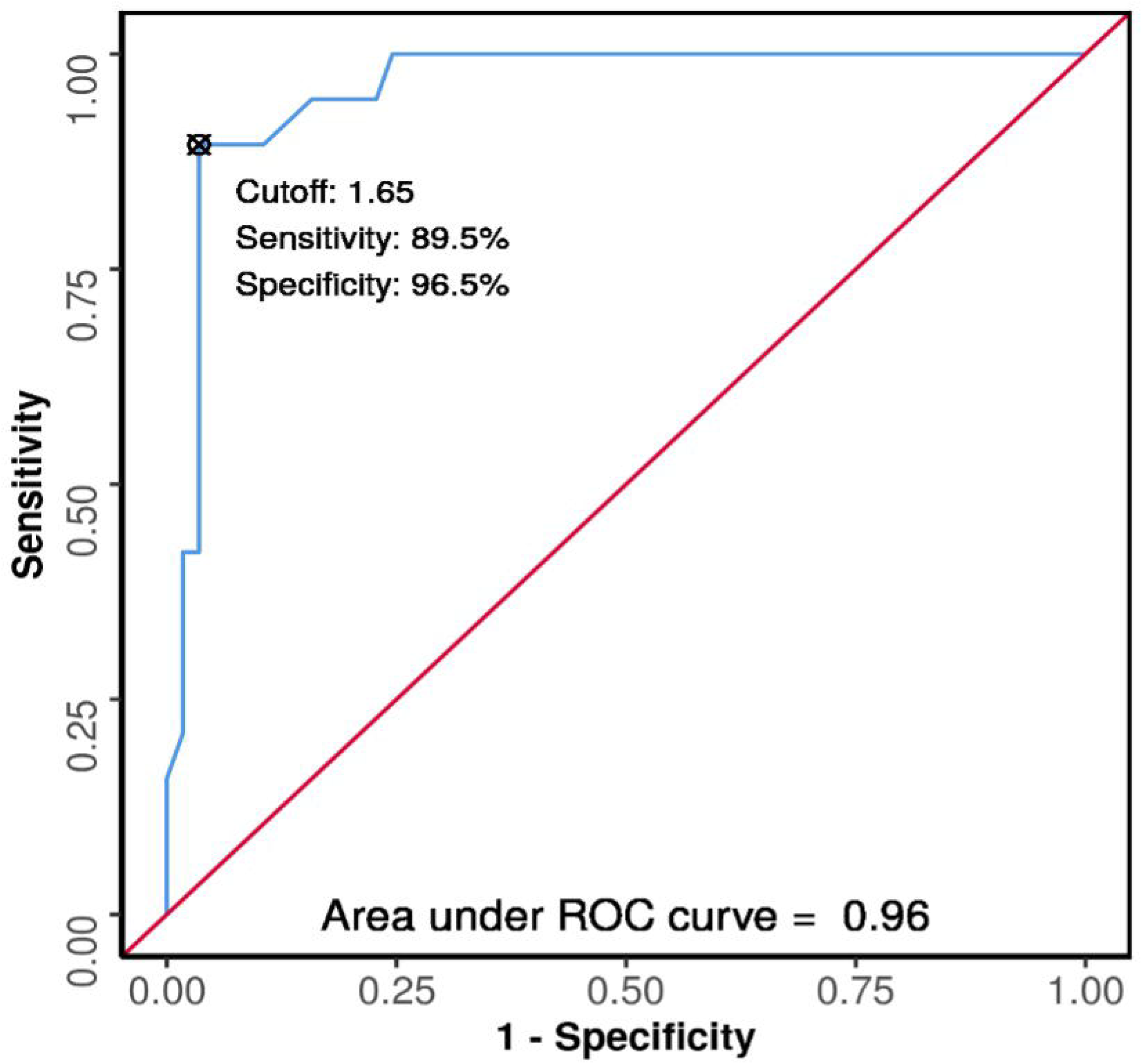

Comparison of QMI was made between patients diagnosed as confirmed sarcopenia using EWGSOP 2 criteria. Group comparison was done by Wilcoxon-Mann-Whitney U Test as the variable QMI was not normally distributed among the groups. There was a significant difference between the 2 groups in terms of QMI (cm/m2) (W = 106.000, p = <0.001), with the median QMI (cm/m2) being highest in the patients of cirrhosis where Sarcopenia (EWGSOP 2) is absent as shown in Table 3 (Supplemental file) and the strength of association (Point Biserial Correlation) =0.56 (large size effect).

An attempt was analyse diagnostic performance of QMI (cm/m2) in predicting sarcopenia using EWGSOP 2 criteria among males and females.

The area under the ROC curve (AUROC) for QMI (cm/m2) predicting Sarcopenia (EWGSOP2) for males with cirrhosis was **0.96** (95% CI: 0.917 - 1) and p= 0.016, thus demonstrating excellent diagnostic performance as shown in Figure 4. At a cutoff of QMI (cm/m^2^) ≤**1.65 cm/m**^**2**^, it predicts Sarcopenia (EWGSOP2) for males with cirrhosis with a sensitivity of 90%, and a specificity of 96%.

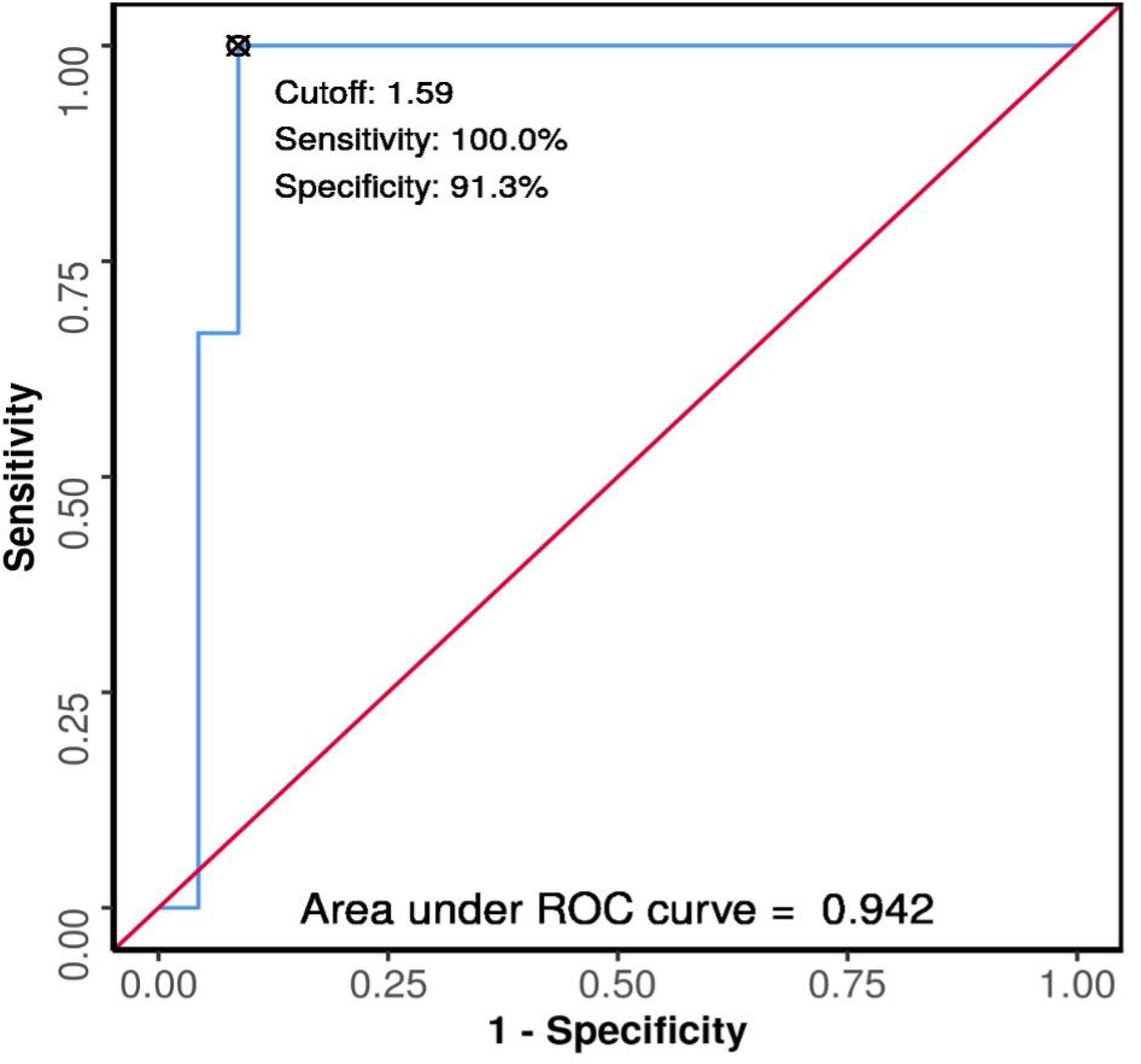

The area under the ROC curve (AUROC) for QMI (cm/m^2^) predicting Sarcopenia (EWGSOP2) in females with cirrhosis was **0.942** (95% CI: 0.849 - 1) and p= 0.016, thus demonstrating excellent diagnostic performance as shown in Figure 5. At a cutoff of QMI (cm/m2) ≤**1.59 cm/m**^**2**^, it predicts Sarcopenia (EWGSOP2) for females with cirrhosis with a sensitivity of 100%, and a specificity of 91%.

## DISCUSSION

The study attempted to evaluate the relationship between QMI and both the MELD score and HGS, which are key clinical indices of liver disease severity and muscular strength. There was a substantial negative association between QMI and MELD score (rho = −0.55, p < 0.001), indicating that lower muscle mass, as measured by QMI, is associated with more severe liver dysfunction.

This finding corroborates existing literature that links sarcopenia with worse liver function and poor prognosis in cirrhotic patients. As cirrhosis progresses, there is often a substantial depletion of skeletal muscle, which further exacerbates the clinical course of liver disease. Thus, detecting sarcopenia at an early stage in these patients enables clinicians to recognize those at higher risk of adverse outcomes and tailor interventions accordingly.

The study also found a substantial positive correlation among QMI and HGS (r = 0.63, p < 0.001). This relationship lends credence to the idea that measuring both muscle mass and strength provides a fuller picture of sarcopenia in cirrhosis patients. While muscle mass assessment helps identify those at risk for sarcopenia, HGS is a functional measure that reflects muscle strength, which is critical for maintaining mobility and independence in cirrhotic patients. Our findings suggest that combining both measures may improve the accuracy of sarcopenia diagnosis at the bedside, allowing for more precise identification of patients requiring intervention.

Additionally, in our study, patients with decompensated cirrhosis showed considerably lower QMI values than those with compensated cirrhosis (Mann-Whitney U = 1172, p = 0.035) further highlighting QMI as a predictor of severity of cirrhosis. Decompensated cirrhosis is frequently linked to declining liver function, and the presence of sarcopenia may further deteriorate the prognosis. These findings highlight the importance of monitoring QMI in decompensated cirrhosis to enable early detection of sarcopenia and timely intervention, helping to prevent its adverse effects on patient outcomes.

These results align with prior studies. **Nevian El-Liethy et al**. performed a study on 101 cirrhotic patients, revealing significant negative correlation between QMI and MELD score (R = −0.532, p < 0.001) and a considerable positive correlation among QMI as well as HGS (R = 0.812, p < 0.001). [8] Similarly, **Mandill et al**. evaluated Quadriceps Muscle Layer Thickness (QMLT) in 93 individuals and found a significant positive correlation between QMLT and HGS (r = 0.423, p = 0.002 in males; r = 0.761, p = 0.001 in females). Furthermore, significant negative correlation between QMLT and Na-MELD scores in men (r = −0.460, p = 0.001) was reported. [10] These observations further support our study’s conclusions, reinforcing the utility of ultrasound-derived muscle measurements in assessing sarcopenia severity and its clinical implications in cirrhotic patients.

The study further identified specific cutoff values for QMI that predict sarcopenia according to the EWGSOP2 criteria, set at ≤ 1.65 cm/m^2^ (AUROC-0.96) for males and ≤ 1.59 cm/m^2^ (AUROC - 0.942) for females. Similar findings were observed by **El-Liethy et al** who identified QMI cutoffs for predicting sarcopenia as 1.67 cm/m^2^ for males (AUROC=0.908) and 1.58 cm/m^2^ (AUROC=0.984) for females. [8]

The study’s findings contribute to the growing evidence supporting ultrasound as a reliable and non-invasive tool for detecting sarcopenia in cirrhosis. Ultrasound offers several advantages, including its widespread availability, low cost, and ease of use, making it an attractive alternative to more expensive and resource-intensive imaging techniques. Furthermore, its ability to assess muscle mass and strength in real-time provides clinicians with a valuable tool for monitoring disease progression and response to treatment.

However, this study does have some limitations. To begin, the study’s cross-sectional design limits its ability to detect cause-and-effect relationships between sarcopenia and liver disease outcomes. A longitudinal study would provide a more complete knowledge of how changes in muscle mass over time affect disease development and clinical outcomes. Second, because this study was carried at a single tertiary care centre, its findings may not be fully generalizable to broader cirrhosis populations. The patient cohort may not represent the full diversity of cirrhosis cases encountered in other healthcare settings, potentially limiting the external validity of the results. Lastly, while ultrasound-based QMI measurement is a safe and convenient approach for evaluating muscle mass, future research comparing it with gold-standard methods such as CT or MRI will be essential to further validate its accuracy and reliability.

## CONCLUSIONS

This study highlights the Quadriceps Muscle Index (QMI), measured via ultrasound, as an effective way for assessing muscle mass and disease severity among cirrhosis patients. The moderate negative correlation between QMI and MELD score underscores its potential in predicting liver disease progression and patient outcomes. Given the rising global burden of liver disease, early identification and intervention for sarcopenia are crucial for improving patient prognosis and quality of life. Future multicentre trials and longitudinal research are needed to enhance the validation of ultrasound-based QMI measurements in routine clinical settings.

## Data Availability

Data will be available on reasonable request

## Acknowledgement

We thank Dr. Shalini Sharma for giving her valuable inputs while editing the draft.

## Author statement

Dr. Sarthak Gaur: Protocol writing, Data collection and analysis, Prepared the final draft for the article and approved it

Dr. Minakshi Dhar: Conceived the idea, protocol design and writing, analysis of data, final preparation of draft and approved it

Dr. Anand Sharma: Data collection, intellectual inputs in the protocol design writing and inputs for final draft of article and approved it.

Dr. Sonal Saran: Data collection, inputs in data analysis and final draft of article and approved it.

Dr. Mukesh Chand Bairwa: Data collection and approved it.

Dr. Monika Pathania: Data collection, analysis of data and final article writing and approved the final draft.

## Data statement

Data will be uploaded on the data repository. Protocol will be uploaded in supplementary files.

## Ethical Approval

Ethical approval was taken from Institutional Ethics Committee (DHR Reg. No.: EC/NEW/Inst/2022/UA/0180) via letter no AIIMS/IEC/23/275 dated 25/08/2023

## Strengths & limitation

- This study provides the valuable insight into the fact that ultrasound can be used as a valuable tool to assess the sarcopenia in patients with cirrhosis.
- Ultrasound being cost effective and easily available modality can help primary care physicians to plan early intervention in these group of patients.
- Cross-sectional design of the study limits its ability to detect cause-and-effect relationships between sarcopenia and liver disease outcomes.

