## Supplementary figures and images for "Co-relation of Quadriceps Muscle index (QMI) with Hand Grip Strength (HGS) and Model for End Stage Liver Disease (MELD) score and cut offs for predicting sarcopenia in patients with cirrhosis: A Cross-Sectional study"

### STUDY FLOW Figure 1.pdf

**STUDY FLOW (Figure 1)**

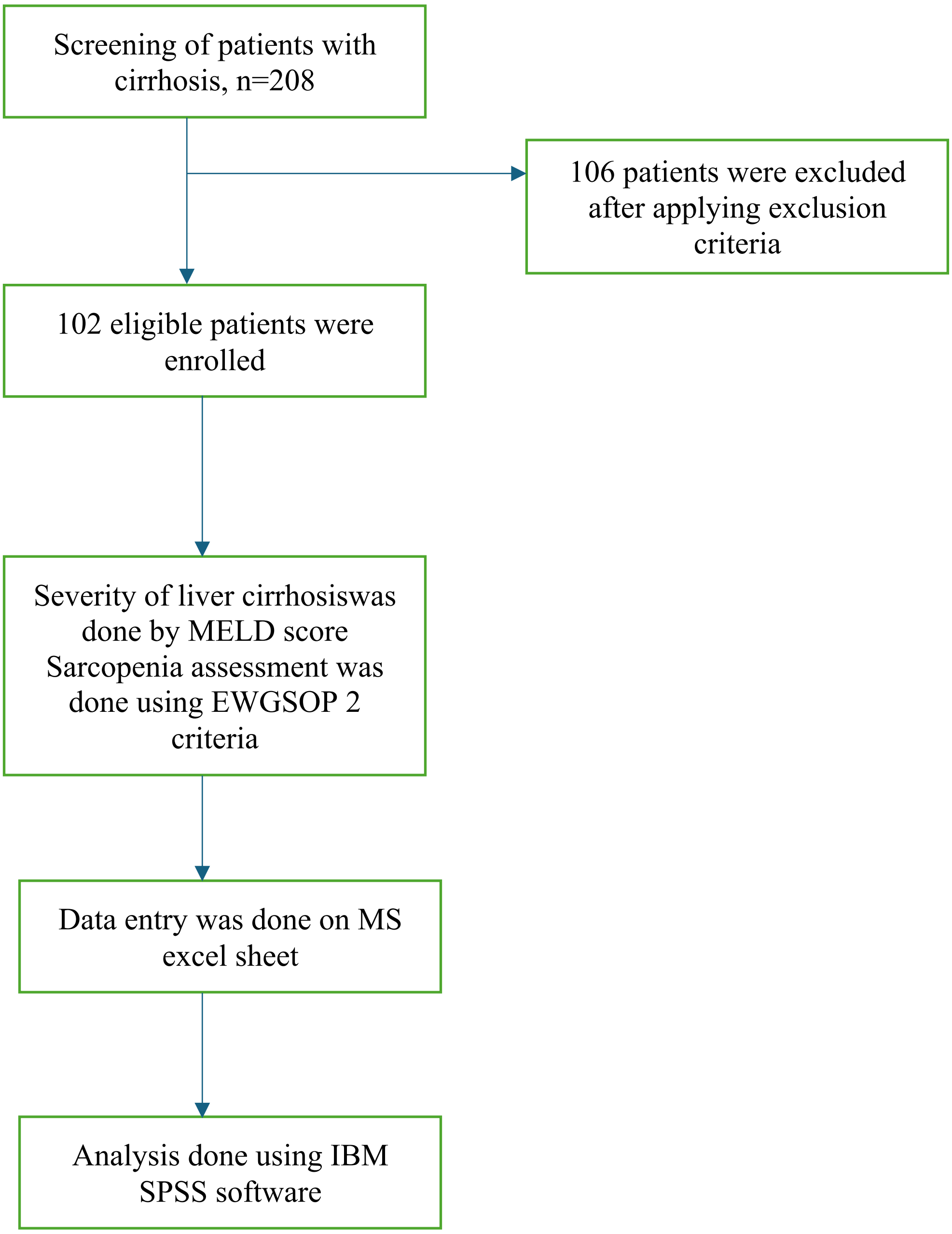
