## Supplemental Table 1 for "Co-relation of Quadriceps Muscle index (QMI) with Hand Grip Strength (HGS) and Model for End Stage Liver Disease (MELD) score and cut offs for predicting sarcopenia in patients with cirrhosis: A Cross-Sectional study"

**Table 1:Patients demographics and Clinical Characteristics**

| **Characteristics** | **Value** |
| --- | --- |
| **Age (Mean ± SD)** | 45.56 ± 11.08 |
| **Age Group (Years) - N (%)** |  |
| 18-30 | 9 (8.8%) |
| 31-40 | 24 (23.5%) |
| 41-50 | 33 (32.4%) |
| 51-60 | 27 (26.5%) |
| 61-70 | 9 (8.8%) |
| **Gender - N (%)** |  |
| Male | 76 (74.5%) |
| Female | 26 (25.5%) |
| **Cirrhosis Type - N (%)** |  |
| Compensated | 23 (22.5%) |
| Decompensated | 79 (77.5%) |
| **MELD Score (Mean ± SD)** | 13.08 ± 3.34 |
| **Anthropometry (Mean ± SD)** |  |
| Height (cm) | Male: 165.87 ± 5.68, Female: 150.08 ± 5.78 |
| Weight (kg) | Male: 64.89 ± 11.50, Female: 54.92 ± 9.19 |
| BMI (kg/m²) | Male: 23.57 ± 3.92, Female: 24.04 ± 3.30 |
| MUAC (cm) | Male: 25.33 ± 3.84, Female: 24.35 ± 3.19 |
| **Muscle Strength & Mass** |  |
| QMI (cm/m²) (Mean ± SD) | 1.82 ± 0.27 |
| HGS (Mean ± SD) | 27.57 ± 7.06 |
