## Supplemental Table 3 for "Co-relation of Quadriceps Muscle index (QMI) with Hand Grip Strength (HGS) and Model for End Stage Liver Disease (MELD) score and cut offs for predicting sarcopenia in patients with cirrhosis: A Cross-Sectional study"

***Table 3: Comparison of QMI between patients with and without sarcopenia as per EWSGOP2 criteria***

| **QMI (cm/m2)** | **Sarcopenia (EWGSOP2)** | | **Wilcoxon-Mann-Whitney U Test** | |
| --- | --- | --- | --- | --- |
|  | **Present** | **Absent** | **W** | **p value** |
| Mean (SD) | 1.53 (0.15) | 1.90 (0.24) | 106.0  00 | <0.001 |
| Median (IQR) | 1.57 (1.46-1.64) | 1.83 (1.73-2.02) |  |  |
| Min - Max | 1.17 - 1.74 | 1.42 - 2.62 |  |  |
